# Detection of a *Vibrio paracholerae* Case in a Diarrheal Disease Outbreak in Costa Rica

**DOI:** 10.64898/2026.05.07.26352676

**Authors:** Estela Cordero-Laurent, Melany Calderon-Osorno, Adriana Godínez-Rojas, Jimena Blanco-Arguedas, Guillermo Barquero-Ureña, Ericka Umaña-Valverde, Gletty Oropeza-Barrios, Grettel Chanto-Chacón, Christine C. Lee, Francisco Javier Duarte-Martinez

## Abstract

First documented detection of *Vibrio paracholerae* in a Costa Rican foodborne outbreak. Genomic analysis confirmed species identity, revealing limitations of conventional PCR and MALDI methods. Findings underscore the need for genomic surveillance to accurately characterize emerging enteropathogens and support public health systems.

## Short communication

At the end of April 2025, the Costa Rican Institute for Research and Teaching in Nutrition and Health (INCIENSA), responsible for laboratory-based epidemiological surveillance in the country (1), responded to an outbreak of acute diarrheal disease at a non-profit foundation located in the province of Alajuela, which serves individuals with severe disabilities and terminal illnesses. According to the national protocol for surveillance of foodborne diseases (2), two clinical stool samples and ten food samples from the food service were received. This activity was reviewed by CDC, deemed not research, and was conducted consistent with applicable federal law and CDC policy^1^.

Rotavirus A was detected in both clinical samples using the BioFire FilmArray (3). Additionally, one sample tested positive by PCR for *V. cholerae* non-O1, non-O139, *ctxA* negative, according to the WHO Global Salm Surv surveillance network (4). In one food sample (vegetable blend), *V. cholerae* was also detected using the BAX Q7 system (5) and the strain was also typed as *V. cholerae* non-O1, non-O139, *ctxA* negative. Both strains were further confirmed by MALDI-TOF (6).

To determine taxonomic identification and confirm genomic relationships between the strains, whole genome sequencing was performed using an Illumina MiSeq system. A 250 cycle paired-end run with a V2 500-cycle flow cell was employed. Libraries were prepared with the Illumina DNA Prep kit, following the standardized procedure of the PulseNet Latin America and Caribbean network(7). *De novo* genome assembly was performed using Shovill, within the pipeline theiaprok_illumina_pe_PHB v3.0.1 on the Terra.Bio platform, which included the following tools: Average Nucleotide Identity (ANI) with mummer version 4.0.0rc1, Gambit-metadata-2.0.0, and Kmerfinder_bacteria_20230911 (8). The analyzed sequences met the quality parameters established by the pipeline. Ribosomal MLST was also utilized for identification (9).

A Neighbor-Joining analysis was conducted to determine phylogenetic relationships between the sequences of the strains and a set of genome sequences from different *Vibrio* species, as described by Islam MT *et al* (10). The reference sequence of *V. paracholerae* (O75TS_SAN5442A2) was also included in the comparative analysis.

The identification tools yielded discordant results. ANI and Kmerfinder classified both strains as *V. cholerae*, while Gambit and rMLST characterized them as *V. paracholerae*. The phylogenetic analysis provided better resolution than the previous tools, showing that the strains clustered within the same monophyletic group of *V. paracholerae* sequences, including the reference sequence O75TS_SAN5442A2. Both strains, food and clinical, were closely related, showing no SNP differences between them. Importantly, they did not belong to the branch of the tree that included *Vibrio cholerae* genomes (see Figure 1).

**Figure 1.**
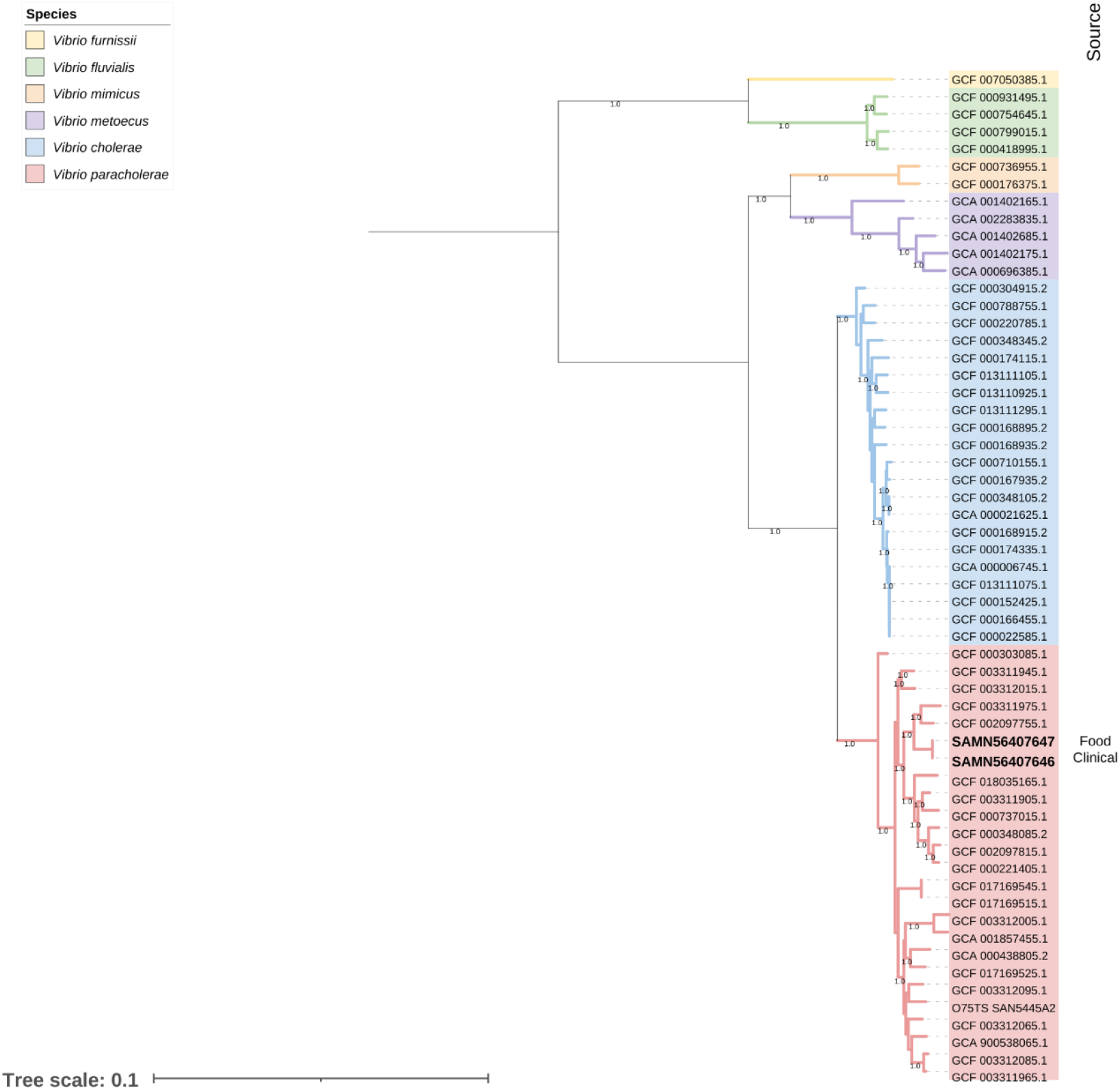
Neighbor-joining analysis for species identification of *Vibrio* sp. Midpoint-rooted phylogenetic tree showing the evolutionary relationships of 56 core genome sequences from different *Vibrio* species and 2 query sequences derived from a clinical and a food sample.

These results were extremely relevant for the epidemiological surveillance system of the country in addition to international reference laboratories that are widely implementing next generation sequencing tools and bioinformatics for outbreak analytics. We documented for the first time the presence of *V. paracholerae* in a foodborne disease event (outbreak) and link to a possible source (vegetable blend). Here, we highlight the limitations of currently available methods (PCR, MALDI, and BAX Q7 system) in discriminating between closely related species (i.e., *cholerae* / *paracholerae)* and emphasize the need for national reference centers to adopt genomic surveillance tools in support of local laboratory networks for proper characterization of persistent and emerging enteropathogens of public health importance.

## Data Availability

All data produced in the present work are contained in the manuscript

## Disclaimer

The findings and conclusions of this report are those of the authors and do not necessarily represent the official position of the Centers for Disease Control (CDC).

## Ethics statement

The data analyzed in this study were generated by the Instituto Costarricense de Investigación y Enseñanza en Nutrición y Salud (INCIENSA) as part of its legal mandate for laboratory-based epidemiological surveillance and specialized public health monitoring, in accordance with the Law No. 4508 (Law of Creation of INCIENSA) and its subsequent amendments, including the Law No. 10805. All laboratory analyses and data generation were conducted as part of routine institutional activities for public health surveillance, under the technical authority of the Ministry of Health of Costa Rica. The study did not involve direct interaction with human subjects or clinical intervention. Data was analyzed in an aggregated and anonymized manner, exclusively for public health and epidemiological purposes. Therefore, in accordance with national regulations this study did not require approval from an institutional research ethics committee.

## Footnotes

1 See 45 C.F.R. part 46

